# Which Measures are Effective in Containing COVID-19? — Empirical Research Based on Prevention and Control Cases in China

**DOI:** 10.1101/2020.03.28.20046110

**Authors:** Shanlang Lin, Junpei Huang, Ziwen He, Dandan Zhan

## Abstract

Various epidemic prevention and control measures aimed at reducing person-to-person contact has paid a certain cost while controlling the epidemic. So accurate evaluation of these measures helps to maximize the effectiveness of prevention and control while minimizing social costs. In this paper, we develop the model in Dirk Brockmann and Dirk Helbing (2013) to theoretically explain the impact mechanism of traffic control and social distancing measures on the spread of the epidemic, and empirically tests the effect of the two measures in China at the present stage using econometric approach. We found that both traffic control and social distancing measures have played a very good role in controlling the development of the epidemic. Nationally, social distancing measures are better than traffic control measures; the two measures are complementary and their combined action will play a better epidemic prevention effect; Traffic control and social distancing do not work everywhere. Traffic control only works in cities with higher GDP per capita and population size, while fails in cities with lower GDP per capita and population size. In cities with lower population size, social distancing becomes inoperative; the rapid and accurate transmission of information, a higher protection awareness of the public, and a stronger confidence of residents in epidemic prevention can promote the realization of the measure effects. The findings above verify the effectiveness and correctness of the measures implemented in China at present, at the same time, we propose that it is necessary to fully consider the respective characteristics of the two measures, cooperating and complementing each other; what’s more, measures should be formulated according to the city’s own situation, achieving precise epidemic prevention; Finally, we should increase the transparency of information, improve protection awareness of the public, guide emotions of the public in a proper way, enhancing public confidence.

## Introduction

Since December 2019, the outbreak of the 2019 novel coronavirus(COVID-19)in Wuhan, China has been spreading rapidly, and now it has been declared a global pandemic by the World Health Organization (WHO). As of 18:00 on March 21st, European Central Time, a total of 267,013 cases have been detected and confirmed worldwide, with a cumulative death of 11,201, and 184 countries and regions with reported cases. Among them, 184,657 cases were confirmed outside China, which is more than twice in China.

Looking back at the outbreak of COVID-19 epidemic and its prevention and control, Wuhan was the first city to report 27 confirmed cases on December 31, 2019, and 554 cases were reported in China by January 22, 2020, of which 444 were in Hubei Province. The week starting on January 24, the Chinese Spring Festival holiday, is the period with the largest passenger flow in a year. In 2019, more than 421 million people were transported by commercial transportation agencies, including trains, cars and aviation, which brings difficulties to COVID-19 prevention and control. On January 23, Wuhan announced the “lockdown” of the city, and the city’s urban bus, subway, ferry, and long-distance passenger transportation were suspended; for no special reason, citizens should not leave Wuhan, and the passages out of Wuhan in the airport and railway station are temporarily closed. On January 23, Zhejiang, Guangdong, and Hunan provinces launched Level Ⅰ emergency response over the Coronavirus. Since January 25, all provinces except Qinghai and Tibet have launched the response. On January 30, Tibet is the last province to launch it. This is the highest-level emergency response adopted in accordance with the *National Public Health Emergency Preparedness* of China, which classifies public health emergencies into particularly major (level I), major (level II), and large (level III) and general (level IV). According to the Level Ⅰ response, the provincial government is responsible for emergency response, adopting prevention and control measures in a timely manner. They adopt isolation, blocking epidemic areas, as long as adopting emergency mobilization and collection of relevant personnel, materials, vehicles, and related facilities and equipment to ensure that all kinds of materials needed for emergency response are sufficient.

The prevention and control measures in China can be summarized into several aspects: the first is to control traffic, which means that vehicles and individuals are not allowed to enter or leave except for the transportation of necessities for urban residents and medical supplies; the second is to stop all or part of bus transportation operations in the city, and there are vehicles and personnel designated by the government to be responsible for the transportation of essential supplies; the third is to stop the operation of non-residential business institutions, including entertainment, sports, public culture and leisure places, and delay the opening of all schools; the fourth is to isolate urban and rural communities, which means non-community residents are not allowed to enter. It also includes performing temperature surveillance on individuals who enters and leaves the community and institutions, and conducting medical examination and screening on the febrile. In hard-hit communities, where residents are not allowed to leave their homes, supplies are pre-ordered online and distributed by community managers and volunteers; the fifth is to trace the confirmed and suspected cases, which means mandatorily isolating and treating the confirmed cases, quarantining the suspected cases centralized, and quarantining the contacts centralized or at home; the sixth is to mobilize medical and protective equipment. Each province sends medical and health personnel and mobilizes medical equipment to support Hubei province, distributes or quantitative sales of masks to residents, and requires residents to wear a face mask when they go out; the seventh is to insist on information disclosure. Seventh, insist on the disclosure of information. From the central government to the local area, regular reports of confirmed and suspected cases of COVID-19 are made daily, and epidemiological follow-up investigation reports are released, including their whereabouts and all possible contacts; the eighth is to quarantine individuals from other cities for 14 days. Since March 14, some areas require overseas entrants to be conducted centralized quarantine for 14 days. January 24 to February 2 is the Chinese Spring Festival holiday. However, after the end of the holiday, enterprises were not allowed to operate. Most provinces stipulated that production operations could only be carried out after February 10 with the approval of the government. They also required that people from other cities and provinces must provide health certificates and be quarantined for 14 days. Specifically, individuals who came from hardest-hit area should be quarantined at a designated location for 14 days, as for those from non-hardest-hit area, they also should be quarantined at home 14 days. After March 20, some provinces will only gradually relax control if there are no new confirmed cases for several consecutive days.

The stringent control measures in China quickly brought the epidemic under control. By February 18, the number of diagnosed cases in Wuhan reached a peak of 38,020, and in other regions reached a peak of 13,886 on February 14, then the number of new cases began to gradually decrease. By March 18, for the first time, there were no new cases within China, and the stock of diagnosed cases fell to 7,263. The prevention and control of COVID-19 have mainly shifted to controlling imported cases overseas. However, of the 32 province-level administrative regions in mainland China, 6 have fewer than 100 confirmed COVID-19 cases, 13 were between 100 - 500 cases and only 5 have more than 1,000 cases, namely Hubei, Guangdong, Henan, Zhejiang, and Hunan. There were no COVID-19 cases in 19 of 334 prefecture-level administrative regions across the country, and more than a third of the 2,851 county-level administrative regions did not occur. In other words, the epidemic in more than half of the country is very mild, while Tibet has only one imported case, but they all have adopted a Level Ⅰ emergency response without exception. The Level Ⅰ emergency response means a suspension of economic activity in all regions of the country, with costs that are difficult to estimate. One percentage reduction in China’s GDP growth rate means a loss of more than 830 billion yuan. At the same time, strict measures to control COVID-19 epidemic will cause economic depression and rising unemployment, as well as increase the number of abnormal death (suicide and malnutrition, insufficient medical care, etc.). Therefore, when the epidemic in China is effectively controlled, it is necessary to reflect on whether it is necessary to implement comprehensive control in regions where there are no infected cases? How much prevention and control is needed to be effective in regions with different epidemic severity?

The main contributions of this article are as follows: First, in response to the control measures of COVID-19 in China, we attempt to use statistical methods to distinguish which measures are effective in different regions, which has rarely been discussed in epidemiology in the past; Second, regarding the differences between measures taken by China in COVID-19 and all the measures in other countries in the past few decades, we take the traffic control measure, the most comprehensive and severe that have not been available in the past few decades, as an independent explanatory variable, trying to analyze which areas need to be adopted. Third, this paper incorporates the proposed effective distance into the model for regression, breaking through the past practice of estimating the scale of epidemic spread based on geographical distance. Fourth, this paper uses Python and other big data mining methods to mine and sort out the prevention and control measures adopted in China and the time taken, as well as the Baidu Search Index “COVID-19”, “the correct wearing of masks”, “Zhong Nanshan” to represent information dissemination, public awareness of protection and Confidence in fighting the epidemic.

### literature review

With the progress of globalization, emerging infectious diseases continue to occur and are more likely to cause a global pandemic. Most of them originate from animals. According to statistics, zoonoses account for up to 60.3% of the new infectious diseases, of which 71.8% originate in wildlife (Jones et. al., 2008). The situation of prevention and control of emerging infectious diseases is very serious (Han et al., 2016). Since the 21st century, emerging infectious diseases have outbroken one after another, such as the SARS in 2003, H1N1 influenza in 2009, H7N9 influenza in 2013, the Ebola hemorrhagic fever in West Africa in 2014, the Zika virus epidemics in South America in 2016 and yellow fever in Angola and Brazil in 2017 are all zoonoses (Zheng et al., 2019), which have posed a serious threat to human life and health and socio-economic development. There are three stages in the pandemic zoonoses emergence: the first stage is the pre-emergence, under which the pathogen is transmitted between their animal reservoirs. The second stage is localized emergence, during which self-limiting spill events or large-scale spills can lead to the person-to-person transmission for a few pathogen generations. The third stage was manifested by widespread and global pandemics. In the first stage, it’s not easy for the prevention and control departments in the government to recognize the characteristics of the epidemic, and they do not understand the infectivity and transmission of the pathogens. Emergency decision-making is a complex process, which means decision-makers must analyze a large amount of uncertain information in a short period of time, and make effective decisions to deal with crises timely. This is an extremely difficult process. Missing that stage, it can form a “butterfly effect”, which causes a huge impact (Morse et al., 2012).

Some literature studies the method that the medical staff treats emerging infectious diseases to reduce or avoid transmission. Koenig (2015) proposed the 3I tool, namely Identify-Isolate-Inform, which is considered to be the normative process for medical staff to treat emerging infectious diseases. The 3I tool has been modified for the prevention and control of infectious diseases such as MERS, measles, Zika virus, mumps, and hepatitis A, and has achieved good results. Triage is an important part of early detection of patients with infectious diseases, effective prevention of the spread of infectious diseases, and better protection of medical staff (Kisting et al., 2012). Medical staff must have a sufficient understanding of emerging infectious diseases. When encountering suspected patients, it’s necessary to ask for medical history in time, to identify potential patients as early as possible, and to make isolation and protection. If a person is suspected to be infected, isolation is required, including the transfer of the patient to another designated place for treatment or direct placement of the patient into an isolation ward. Isolation is an important measure, but there are differences between isolation and quarantine. Quarantine refers to the separation of individuals who may have been exposed to an infectious disease or pathogens but still healthy from those who are healthy and have not been exposed (Chea et al., 2014). In other words, quarantine is to conduct observation on healthy people who may have been exposed to infectious diseases and may spread the disease to others to determine whether they are truly infected. The concept of observation originated from “border screening”(Selvey et al., 2015). It was originally used for the entry and exit ports, and then expanded the concept to refer to measures taken by people who may be exposed to the disease before symptoms are identified, which are quarantine measures and have no place limitations. For example, people who do not need to be hospitalized can be quarantined at home while being assessed for MERS infection Koenig et al., 2017). During the observation, once symptoms are identified and the infection is confirmed, quarantine should be switched to isolation. Diseases listed for quarantine must be diseases that pose a serious threat to public health, such as cholera, diphtheria, infectious tuberculosis, plague, smallpox, yellow fever, viral hemorrhagic fever, SARS, and influenza which may cause a pandemic (Koenig, 2012). Measures to be taken is determined according to the quarantine and isolation decision tree, and a certain risk-benefit analysis is required. If the symptoms are not contagious before onset, the travel and social distance will not be limited, but necessary disease surveillance should be carried out to determine whether the intervention of public health agencies is needed to ensure the effectiveness of the latent disease surveillance based on the monitoring effect. Different public health measures need to be selected based on the disease to which the individual assessed is exposed. On the other hand, the difference transmission can significantly affect the speed of disease spreading and the choice of personal protective equipment (PPE). At the same time, legal basis, jurisdiction, impact on mental health and the potential negative consequences of restrictions on human rights and free actions need to be considered. The scientific significance of quarantine lies in the fact that quarantined disease is likely to spread during the incubation period to minimize the probability of infectious disease outbreaks (Barbisch et al., 2015). The third step of the 3I tool is to promptly report to the infection control department of hospitals, superior supervisors, and law enforcement departments based on the location of the identified and confirmed patient.

Another part of the literature assesses the effectiveness and the scope of application of public health policies. Rothstein & Talbott (2007) found that to make quarantine to be the most effective measure for limiting the spread of infection, compliance with quarantine regulations should be encouraged, that is, providing job security and income replacement. Wang et al. (2012) found that In the case of short reaction time, intra-population, such as patient isolation, are more effective than inter-population such as travel restrictions. Jefferson (2008) investigated the effectiveness of physical interventions to interrupt or reduce the spread of respiratory viruses. Experiment result showed that many simple and low-cost interventions, such as washing hands, wearing a mask and isolating patients may be infected, can effectively prevent the spread of respiratory virus infections. Physical measures are very effective in preventing the spread of SARS. The incremental effect of adding a viricide or preservative to routine hand washing to reduce the spread of respiratory disease remains uncertain. The effect of screening and social distancing at ports of entry cannot draw a clear conclusion. Sakaguchi et al. (2012) tested border Control measures and community containment in Japan during the early stages of 2009 pandemic influenza (H1N1) and found those were effective to some extent. Rashid (2015) researched the public health policy of the 2009 pandemic influenza and concluded that the school closure, whether positive or passive, seemed to be moderately effective and acceptable in reducing the spread of influenza and delaying the peak of the epidemic, but it is related to high expenses of secondary school. Voluntary isolation and quarantine at home are also effective and acceptable measures, but it takes the risk of intra-household transmission from index cases to contacts. Workplace-related interventions, such as work closure and work at home, are also moderately effective and acceptable, but it may have negatively impact on economy. Internal mobility restriction is only effective when extremely high restrictions (50% of travel) are imposed and mass gatherings which happened within 10 days before the epidemic peak is likely to increase the risk of transmission of influenzas. Rothstein (2015) compared the effectiveness of public health policies to intervene with infectious diseases, among which quarantine is one of the most aggressive and controversial measures that public health officials can take when trying to control an outbreak. Serious legal and ethical concerns have been raised because of restrictions on the movement of potentially large numbers of people (Fidler et al., 2007). During the SARS epidemic in 2003, quarantine was widely used in several Asian countries and Canada, and it played a role in ending the epidemic. Similarly, during the 2014 Ebola outbreak in West Africa, social distancing measures, including quarantine, had become a major containment strategy. The paradox of quarantine and other social distancing measures is that they may effectively resist the outbreak of disease, but the scope of application is too wide, leading to various social hazards, including economic disruption, physical isolation and even violence. Quarantine is the first public health tool to control infectious diseases and remains a legitimate and valuable public health strategy because it has a significant impact on civil liberties, economic activity and social cohesion, so officials must be extra careful about the possibility of abusing quarantine authority(Rothstein, 2015).

Huremović(2019) analyzed and compared the clear definitions and scope of common public health policies, such as isolation, quarantine, shelter-in-place, cordon sanitaire, protective sequestration, and social distancing. Isolation is to restrict the persons who have an infectious disease in a specified geographical area, separating them from healthy people and those who have not been exposed. Different from isolation, quarantine is to separate those who are still healthy but possibly exposed to an infective agent from those who are healthy but have not been exposed, which is a restriction on the movement or communication of persons or the transport of goods in order to prevent the spread of disease (Gostin & Wiley, 2016). Shelter-in-place (SIP) is a variant of quarantine, in which individuals are not restricted at a designated location, but at where they were at the time (e.g. Family). Cordon sanitaire refers to restricting activities of the individual in a larger and more defined geographic area (such as a community). The “Cordon sanitaire” may be considered a reasonable and useful measure in which: (1) the infection is highly virulent (infectious and possibly cause disease); (2) the mortality rate is extremely high;(3) treatment is not available or difficult to conduct;(4) there is no vaccine or other method to immunize a large number of people. In addition to legal and ethical challenges, the measure is also related to challenges in the logistical resources. Comparing with quarantining at a designated place, the advantage of cordon sanitaire is that it tends to keep the community intact, including its basic resources, businesses, and infrastructure. A method that is the opposite of it is called protective sequestration, which means that unaffected communities separate themselves from the outside until there is no risk of an outbreak. Social distancing is a voluntary, recommended restriction on physical contacts, such as staying at home and avoiding public places and activities. Some less drastic, but quite effective and more frequently used measures of social distancing include canceling mass gatherings, school, and workplace closures, travel restrictions, etc. Wilder-Smith (2020) believed that SARS-CoV has now been eliminated, indicating that we can completely stop this epidemic and interrupt person-to-person transmission. In the absence of vaccines and antivirals, only strict implementation of traditional public health measures made this remarkable achievement possible. In the early stages, isolating patients is particularly effective in interrupting the spread of infectious diseases. Since the infected can spread before he is symptomatic, isolation is often too late to prevent transmission and contain the pandemic. Quarantine is one of the oldest and most effective tools for controlling outbreaks of infectious diseases (Cetron & Landwirth, 2005), which possibly apply at the individual or group level and usually involves restrictions on houses or designated facilities. Social distancing aims to reduce interactions between people in the wider community, where individuals may be contagious but have not yet been identified and therefore have not been isolated. Social distancing is particularly useful where community transmission is considered to be occurring, but the links between cases are unclear, or where restrictions on the known contacts are not considered sufficient to prevent further spread. Social distancing includes closing schools or office buildings, suspending public markets, and canceling gathering. If these measures are considered inadequate, then a community-wide containment is likely to be required. A community-wide containment is an intervention that applies to an entire community, city, or region, aiming to reduce physical contact between people. Implementing a community-wide containment requires close partnerships and cooperation with law enforcement agencies at the local and state levels, and inspection stations are usually established, and legal penalties may be required if there is a violation.

Some scholars studied the effects of travel restrictions. Chong & Zee (2012) found that antiviral drugs and hospitalization were more effective in reducing morbidity than travel restrictions. As for air travel, it’s necessary to target at the main route to the source of influenza, and if 99% air travel restriction is implemented, the peak of the epidemic will be postponed for up to two weeks. Mao (2013) believed that it is not cost-effective to use travel restriction alone, but if 50% of travel restrictions are implemented, combined with antiviral prevention and workplace closure, it is the best strategy. For large urbanized areas, closing the affected workplaces can be an effective and cost-saving strategy to reduce flu outbreaks and it is also the most cost-effective.

Folayan & Brown (2015) found that in order to control Ebola, more than 50 countries have issued travel restrictions on affected countries. However, travel restrictions have made it more difficult to tackle the disease, which is not a solution to contain Ebola, because it has become more difficult to deliver supplies, equipment and humanitarian assistance to the affected areas. When a suspected traveler from West Africa who are symptomatic is quarantined, the isolation facilities must be optimal, which means providing physical, social and psychological support for all persons quarantined and quality personal protective equipment must be provided for health care providers

Although there are lots of searches on public health policies to deal with infectious diseases. However, the prevention and control measures against COVID-19 taken in China are unprecedented, which have been beyond the scope of study in existing literature, such as implementing traffic restrictions across the country and closing communities on a large scale. Therefore, it is still necessary to classify the prevention and control measures of China and study which measures are more effective in different regions.

### Theoretical Framework

The outbreak of infectious diseases is a complex process of spreading that occurs in a population. There is a long history of modeling infectious disease epidemics (Anderson et al., 1992), and various modeling specifications have been developed. Kermark & Mckendrick (1927) proposed the classic SIR model, which is a compartmental model. In the SIR model, it is assumed that each individual is the same, the population is homogeneous mixing, the contact is instant and independent of history, infection rate, recovery rate is constant. Individuals in the same state form a compartment, and as the state changes, they move among the compartment.

With the growth of urban population and the development of transportation networks, social mobility has increased, and the spatial expansion of infectious diseases has shown a new pattern, especially when people move among different regions, the spread of the epidemic is common. Understanding the impact of population flow patterns on the prevalence of infectious diseases has attracted considerable attention (Gonzalez et al., 2008), and a meta-population model derived from ecology has been applied in the field of the epidemic. With the rise of complexity science, the micro-modeling specification has been developed and combined with social networks, a network-based micro-individual modeling method has been proposed, which provides a new approach to understand the spread of infectious diseases. These models enable epidemiologists and health authorities to understand the transmission process, predict its impact on healthy populations, and assess the effectiveness of different mitigation and prevention strategies. But, in any case, the epidemic of infectious diseases is inseparable from two key factors, namely, person-to-person contact and population mobility. When infectious disease outbreaks, various countries adopt public health intervention policies, in which one is to control human contacts, such as isolation, quarantine, and social distancing; the other is to control population mobility, such as travel restrictions.

The factors affecting the spread of the epidemic are extremely complex, which is closely related to the carrier, environment, weather, the contact and so on. If a city is abstracted into a node beyond the scale of micro-individuals, the population flow between cities is the key factor affecting the scale of virus transmission among cities, which is related to urban density and transportation. The meta-population model is a commonly used model for analyzing the spread of epidemics, which nodes are used to represent the city, and the linings are used to represent the traffic between cities, such as the plane, high-speed rail, bus and so on. Within each node, a certain number of people are set. Colizza et al. (2007) used a meta-population model to characterize human behavior on a global scale. They found that a large-scale epidemic would only erupt when the population density exceeded a certain threshold. When the scale of the network is infinite, a few large nodes (namely, important cities) in the network will cause an epidemic to outbreak globally. Regarding transportation, Brockmann & Helbing (2013) found that the spread of disease is closely related to the “effective distance” between cities rather than geographical distance. If the probabilistically motivated effective distance is used instead of conventional geographic distances, then complex spatiotemporal patterns can be reduced to surprisingly simple, homogeneous wave propagation patterns. It can also reliably predict the arrival time of the disease, and even if epidemiological parameters are unknown, we can still get the relative arrival times and the spatial origin of the propagation process by the approach.

The effective distance is defined as the best mode of transportation between two cities, depending on the probabilistic traffic flow. Assuming that *p*_*mn*_ is the conditional probability of the connectivity matrix 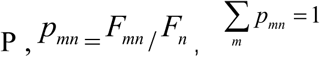。 Weighted links *F*_*mn*_ quantify direct air traffic (passengers per day) from node m to node n, 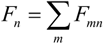 Then,the relationship between effective and *p*_*mn*_ is given as:

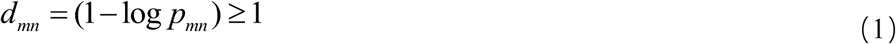

Based on the SIR model, Brockmann & Helbing (2013) proposed an epidemic diffusion model on transportation network, which the equations are as follows:

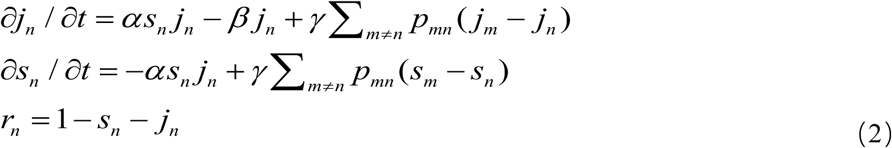

where, *j*_*m*_ and *j*_*n*_ are the local fractions of the infected of city m and city n, *s*_*m*_ and *s*_*n*_ are the local fractions of the susceptible, *r*_*n*_ is recovered population of city n.*α* 、*β* 、*γ* are parameters.

In order to analyze the effect when adopting public health policies to intervene in the spread of infectious diseases, referring to the approach in Brockmann & Helbing (2013) and Zhang (2020), we introduce a term of the effective invasion threshold *ζ* :

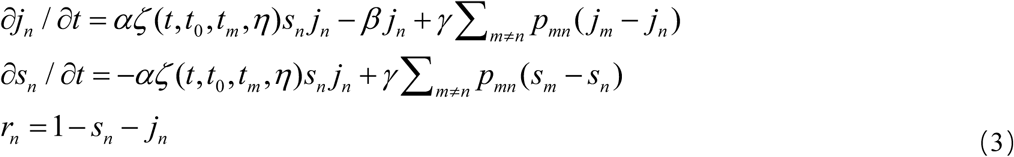

Where, *ζ* is the human intervention factor of public health policy, *η* is the minimum intensity of intervention, *t*_0_ is the time point of human intervention, *t*_*m*_ is the point at which an infectious disease is completely eliminated。Therefore, *ζ* is a function of time and human intervention.

According to equation (3), the scale and speed of COVID-19 transmission are related to factors such as infection rate, recovery rate, and migration rate, as well as effective distance and public health policy intervention. Any public health policy intervention requires costs, and the cost of different interventions varies. In this paper, the various measures to prevent and control the spread of COVID - 19 in China are divided into two categories: One is traffic control, which not only affects the movement of people across regions and even across the country, but also affects the transportation of goods. The price of this is to force companies and other institutions to stop operations, which is at huge cost. Another is social distancing, including closure of public places in the city, community containment, isolation of the infected, and quarantine of the suspected, which affects the operation of some urban institutions, and the cost is less than the traffic control. Based on the mechanism analyzed above and the classification of prevention and control measures against COVID-19, we then propose the ***hypothesis***: traffic control and social distancing can quickly contain the spreads of COVID-19 epidemic, but they perform different effect in various regions.

According to *National Public Health Emergency Preparedness* of China, the main body launching the first-level response is the provincial government, the measures and time adopted by different regions are not the same, and the intensity of implementation also varies from city to city, which provides a good condition for the research in this article.

### Empirical Specification

#### 4.1 Empirical Model and Variable

According to the classification of prevention and control measures of COVID - 19 in China, the baseline estimation strategy is as follows:

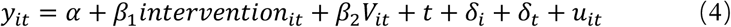

Where, *y*_*it*_is the is the actual cumulative case growth rate of city i in year t. Given that average incubation period of COVID-19 is 5.2 days (Li, 2020), we use the 5th lead of the reported cumulative case growth rate to proxy the actual cumulative case growth rate. *intervention*_*it*_is the main explanatory variable,namely traffic control and social distancing.

*β*_1_ is the coefficient estimated, if *β*_1_ < 0, then it indicates that the prevention and control measures have effectively reduced the growth of cumulative cases and mitigated the outbreak of the epidemic. *V*_*it*_ is control variable, including population size (pop), GDP per capita (pergdp), number of medical institutions (hospital), number of beds in medical institutions (bed), effective distance from Wuhan (distance) to control the city characteristics on the spread of the epidemic. t is time trend,*δ*_*i*_is a region fixed effect to control the characteristics of provinces constant over time, *δ*_*t*_ is a time fixed effect to control to control the time factors that do not vary from individual to individual.

### 4.2 Data

#### a. Construction of the Measure Scoring Indicator

Traffic control and social distancing in each city are classified and scored according to the preventive and control measures taken by prefecture-level administrative regions in China. There are divided into 15 items (see Table 1), each with a score of 1. Scoring starts until the measure is canceled. For example, on January 21, Shanghai began to implement “quarantining the contacts for 14 days.” Since this measure is under “social distancing”, then the score of social distancing in Shanghai from January 21 was 1. On January 24, Shanghai began to implement “closing part of the indoor urban public places”, since this measure is also under “social distancing”, then “social distancing” in Shanghai will be added another 1 point since January 24, and so on, and finally, the points for traffic control and social distancing will be added up.

**Table 1.**
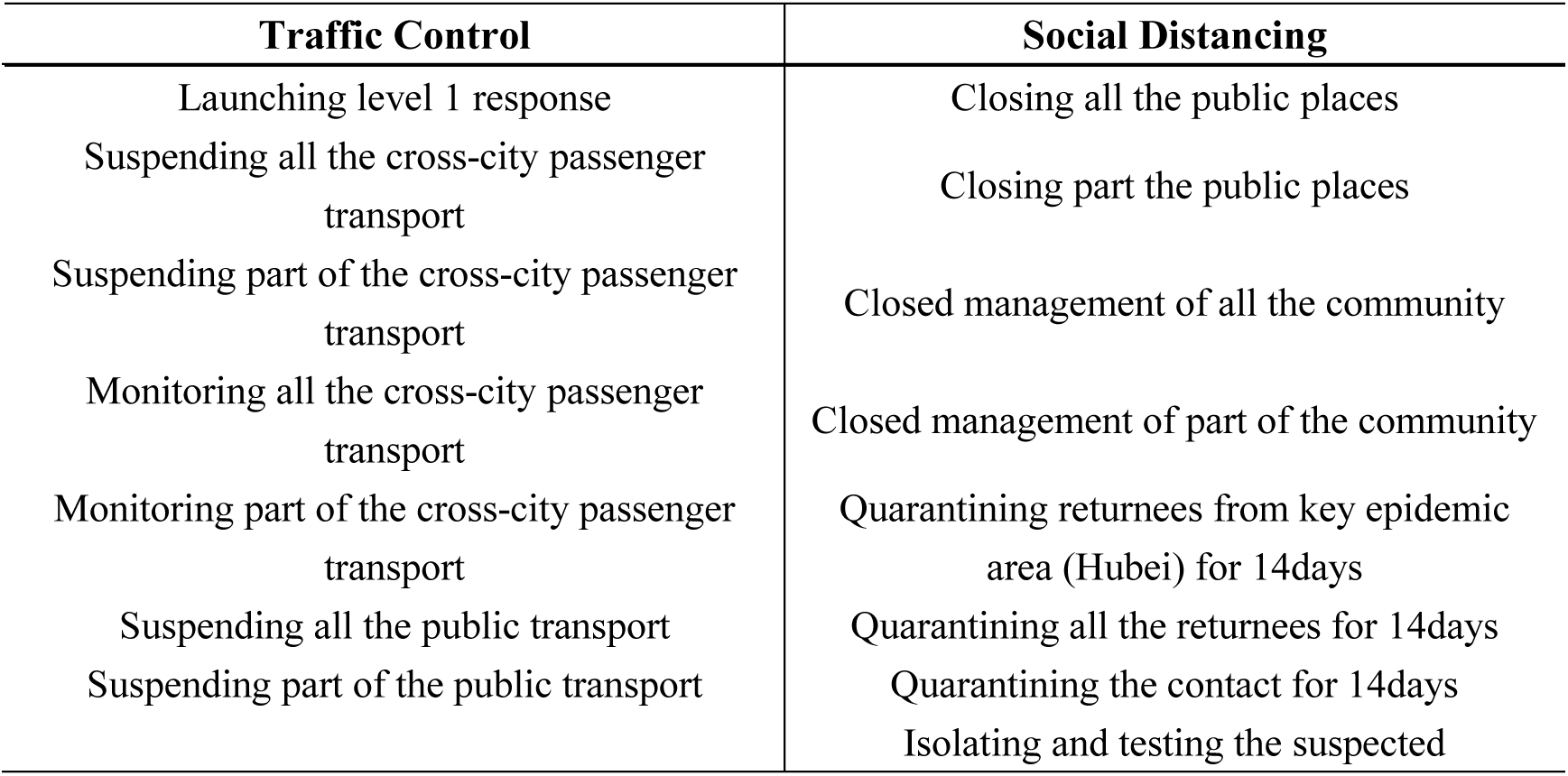
Points for traffic control and social distancing

#### b. Data Source

As to the data used in this paper, the specific content and implementation time of the prevention and control measures came from the information or announcements issued by the prevention and control headquarters of the prefecture-level administrative districts; the cumulative confirmed case of came from the official release of the *National Health and Health Commission*; population size, GDP per capita, number of medical institutions, and number of beds in medical institutions are from the *China City Statistical Yearbook*. Baidu search index data comes from the *Baidu Index* website. We refer to the approach of Brockmann and Helbing (2013) to calculate the effective distance from Wuhan to each city. The inter-city passenger flow data used in the calculation are from *Baidu Migration* website. The time span of the data we use is from January 1, 2020 to February 10, 2020. The explanation of each variable is shown in Table 2.

**Table 2.** Variable Explanation

| Attribute | Name | Explanation |
| --- | --- | --- |
| explained variable | Rate | actual cumulative case growth rate |
| explanatory variable | Total Score | Total score of prevention and control measures |
|  | Traffic Control | score of traffic control measures |
|  | social distancing | score of social distancing measures |
|  | Total Score_ dummy | Dummy variable of whether high total score |
|  | Traffic Control_ dummy<br>(used in robustness) | Dummy variable of whether high traffic control score |
|  | Social Distancing_ dummy<br>(used in robustness) | Dummy variable of whether high social distancing score |
| control variable | Pergdp | GDP per capita |
|  | Pop | Population size |
|  | Hospital | Numbers of medical institutions |
|  | Bed | Numbers of beds in medical institutions |
|  | Distance | Effective distance |
| classified variable | Information | Baidu Index of “COVID-19” |
|  | Awareness | Baidu Index of “the correct way to wear a mask” |
|  | Confidence | Baidu Index of “Zhong Nanshan” |

The data samples in this paper consist of the balance panel data of 279 prefecture-level cities from January 1 to February 10, 2020, and the descriptive statistics of related variables are shown as follows.

## Result

### 5.1 Are They Effective?

#### 5.1.1 Baseline Regression

Table 4 reports the baseline regression results estimated using the LSDV (Least Square Dummy Variables) method. The explanatory variable is the actual cumulative cases growth rate. The explanatory variables in column (1) and column (2) are the total score total score of prevention and control measures, in column (3) and column (4) are the score of traffic control measures, in column (5) and column (6) are the score of social distancing. Column (2), (4), and (6) introduce control variables on the basis of columns (1), (3), and (5). The regression results show that the coefficients of the total score, traffic control, and social distancing are significantly negative, regardless of whether control variables are added, which indicates that the total effect of the two measures, traffic control and social distancing alone all significantly reduce the cumulative case growth rate, effectively containing the spread of the epidemic. In addition, the coefficient of social distancing is larger, indicating that the effect of social distancing measures are better than that of traffic control.

**Table 3.** Statistical Description

| VARIABLES | N | mean | sd | min | max |
| --- | --- | --- | --- | --- | --- |
| Rate | 13,950 | 0.0848 | 0.443 | 0 | 19 |
| Total Score | 13,950 | 3.729 | 3.912 | 0 | 10 |
| Traffic Control | 13,950 | 1.758 | 1.802 | 0 | 4 |
| social distancing | 13,950 | 1.972 | 2.252 | 0 | 6 |
| Total Score_dummy | 13,950 | 0.486 | 0.500 | 0 | 1 |
| Traffic Control_dummy | 13,950 | 0.454 | 0.498 | 0 | 1 |
| Social Distancing_dummy | 13,950 | 0.470 | 0.499 | 0 | 1 |
| Pergdp | 13,950 | 92,348 | 379,890 | 17,890 | 6.400e+06 |
| Pop | 13,950 | 171.3 | 226.3 | 16 | 2,451 |
| Hospital | 13,950 | 86.89 | 108.8 | 11 | 1,115 |
| Bed | 13,950 | 12,906 | 17,135 | 920 | 142,708 |
| Distance | 13,950 | 4.100 | 0.678 | 1.883 | 5.707 |
| Information | 13,950 | 179.5 | 287.4 | 0 | 4,655 |
| Awareness | 13,950 | 2,893 | 5,737 | 0 | 84,380 |
| Confidence | 13,950 | 893.4 | 2,172 | 0 | 74,929 |

**Table 4.** Baseline Regression

|  | (1) | (2) | (3) | (4) | (5) | (6) |
| --- | --- | --- | --- | --- | --- | --- |
| Total Score | -0.0239***<br>(0.0030) | -0.0248***<br>(0.0030) |  |  |  |  |
| Traffic Control |  |  | -0.0260***<br>(0.0065) | -0.0257***<br>(0.0065) |  |  |
| Social Distancing |  |  |  |  | -0.0342***<br>(0.0040) | -0.0360***<br>(0.0040) |
| Constant | -0.0961*** | -0.3725** | -0.0708*** | -0.3289** | -0.0958*** | -0.3884** |

|  | (0.0133) | (0.1554) | (0.0129) | (0.1557) | (0.0132) | (0.1554) |
| --- | --- | --- | --- | --- | --- | --- |
| Observations | 12,555 | 12,555 | 12,555 | 12,555 | 12,555 | 12,555 |
| R-squared | 0.031 | 0.038 | 0.025 | 0.034 | 0.026 | 0.039 |
| Control Variables | NO | YES | NO | YES | NO | YES |
| Time Trend | YES | YES | YES | YES | YES | YES |
| Province FE | YES | YES | YES | YES | YES | YES |
| Time FE | YES | YES | YES | YES | YES | YES |

#### 5.1.2 Robustness

In this section, we conduct robustness test from the following two aspects:

①Considering the earliest outbreak was in Hubei province, which is also the first province to lockdown the city, we exclude Hubei Province and re-estimates the coefficient. ② we change the identification strategy, using DID (Difference in differences) approach to assess the effectiveness of the policy. We convert the total score, score of traffic control and social distancing into dummy variable of whether it is high total score (Total Score_dummy), dummy variable of whether it is high traffic control score (Traffic Control_dummy) and dummy variable of whether it is high social distancing score (Social Distancing _dummy), and construct a generalized DID model.

Table 5 panel A reports the result of subsample without Hubei province, and panel B reports the result of DID approach. It can be seen that all the coefficients are significantly negative, and the conclusion is still robust after changing the sample selection and identification strategy.

**Table 5.** Robustness

| Panel A without Hubei Province |  |  |  |  |  |  |
| --- | --- | --- | --- | --- | --- | --- |
|  | (1) | (2) | (3) | (4) | (5) | (6) |
| Total Score | -0.0208***<br>(0.0023) | -0.0215***<br>(0.0023) |  |  |  |  |
| Traffic Control |  |  | -0.0176***<br>(0.0053) | -0.0170***<br>(0.0053) |  |  |
| Social Distancing |  |  |  |  | -0.0314***<br>(0.0031) | -0.0330***<br>(0.0031) |
| Constant | -0.0839***<br>(0.0105) | -0.2589**<br>(0.1283) | -0.0589***<br>(0.0102) | -0.2211*<br>(0.1286) | -0.0861***<br>(0.0105) | -0.2768**<br>(0.1282) |
| Observations | 12,015 | 12,015 | 12,015 | 12,015 | 12,015 | 12,015 |
| R-squared | 0.039 | 0.043 | 0.034 | 0.037 | 0.041 | 0.045 |
| Control Variables | NO | YES | NO | YES | NO | YES |
| Time Trend | YES | YES | YES | YES | YES | YES |
| Province FE | YES | YES | YES | YES | YES | YES |
| Time FE | YES | YES | YES | YES | YES | YES |
| Panel B DID approach |  |  |  |  |  |  |
|  | (1) | (2) | (3) | (4) | (5) | (6) |
| Total Score_ dummy | -0.0898***<br>(0.0189) | -0.0909***<br>(0.0189) |  |  |  |  |
| Traffic Control_dummy |  |  | -0.1891***<br>(0.0154) | -0.1893***<br>(0.0154) |  |  |
| Social Distancing<br>_dummy |  |  |  |  | -0.0757***<br>(0.0164) | -0.0809***<br>(0.0164) |
| Constant | -0.0710***<br>(0.0128) | -0.3284**<br>(0.1556) | -0.0140<br>(0.0121) | -0.2414<br>(0.1564) | -0.0728***<br>(0.0129) | -0.3466**<br>(0.1557) |
| Observations | 12,555 | 12,555 | 12,555 | 12,555 | 12,555 | 12,555 |
| R-squared | 0.032 | 0.035 | 0.021 | 0.023 | 0.032 | 0.035 |
| Control Variables | NO | YES | NO | YES | NO | YES |
| Time Trend | YES | YES | YES | YES | YES | YES |
| Province FE | YES | YES | YES | YES | YES | YES |
| Time FE | YES | YES | YES | YES | YES | YES |

#### 5.1.3 Instrumental Variable

Since the spread of the epidemic may be affected by some unobservable factors, the problem of omitting variables may not be avoided in the regression. At the same time, the places with severe epidemic situation are more inclined to take more vigorous prevention and control measures, that is to say, the development of the epidemic situation will also reversely affect the local prevention and control efforts, so there is a problem of reverse causality. In order to address the endogenous problems caused by missing variables and reverse causality, we use the instrumental variable approach to re-estimate. The selection of instrument variables needs to meet two requirements: the first is that there is a significant relationship between the instrumental variables and the endogenous explanatory variables; and the second is that the instrument variable must be exogenous. We take the 1st lag of the explanatory variable as a instrument variable, and adopt the two stage least squares method (2SLS) and the generalized method of moment (GMM) to estimate it. Table 6 reports the estimated results of the instrumental variable approach. We can say that there is no obvious change in the sign and significance of the coefficients. The results of the instrumental variables approach are consistent with the baseline regression, indicating that the conclusion is still valid after considering endogeneity. Both the traffic control and social distancing have been effective in controlling the epidemic.

**Table 6.** Instrumental Variable

|  | (1) | (2) | (3) | (4) | (5) | (6) |
| --- | --- | --- | --- | --- | --- | --- |
|  | 2SLS | GMM | 2SLS | GMM | 2SLS | GMM |
| Total Score | -0.0361***<br>(0.0034) | -0.0361***<br>(0.0032) |  |  |  |  |
| Traffic Control |  |  | -0.0562***<br>(0.0079) | -0.0562***<br>(0.0086) |  |  |
| Social Distancing |  |  |  |  | -0.0457***<br>(0.0045) | -0.0457***<br>(0.0033) |
| Constant | -0.4647***<br>(0.1718) | -0.4647***<br>(0.1691) | -0.3952**<br>(0.1722) | -0.3952**<br>(0.1674) | -0.4913***<br>(0.1718) | -0.4913***<br>(0.1689) |
| Observations | 12,276 | 12,276 | 12,276 | 12,276 | 12,276 | 12,276 |
| R-squared | 0.033 | 0.033 | 0.026 | 0.026 | 0.029 | 0.029 |
| Control Variables | Yes | Yes | Yes | Yes | Yes | Yes |
| Time Trend | YES | YES | YES | YES | YES | YES |
| Province FE | YES | YES | YES | YES | YES | YES |
| Time FE | YES | YES | YES | YES | YES | YES |

### 5.2 Are They Complementary?

We have proven the effectiveness of the two prevention and control measures. Is there a complementary relationship between traffic control and social distancing? We introduced both the two variables into the empirical model for a further analysis, which is shown in Table 7 reports. Column (1) and column (2) are the results of the score data, while column (3) and column (4) are the estimated results of the DID with dummy variables. The results show that the interaction term is significantly negative and has the same sign as the main explanatory variable, that is, the two measures show a complementary relationship, which implies that the combined effect of the two types of measures can more effectively prevent the further spread of the epidemic.

**Table 7.** the Complementary Relationship between Traffic control and Social Distancing

|  | (1) | (2) | (3) | (4) |
| --- | --- | --- | --- | --- |
| Traffic Control_score | 0.0151*<br>(0.0078) | 0.0160**<br>(0.0078) |  |  |
| Social Distancing_score | 0.0158*<br>(0.0092) | 0.0123<br>(0.0092) |  |  |
| Traffic Control_dummy |  |  | 0.0722*** | 0.0737*** |
|  |  |  | (0.0256) | (0.0256) |
| social distancing _dummy |  |  | 0.0715** | 0.0612* |
|  |  |  | (0.0334) | (0.0334) |
| Interaction terms_score | -0.0147*** | -0.0143*** |  |  |
|  | (0.0025) | (0.0025) |  |  |
| Interaction terms_dummy |  |  | -0.2010*** | -0.1954*** |
|  |  |  | (0.0377) | (0.0377) |
| Constant | -0.0904*** | -0.3751** | -0.0702*** | -0.3510** |
|  | (0.0134) | (0.1553) | (0.0129) | (0.1556) |
| Observations | 12,555 | 12,555 | 12,555 | 12,555 |
| R-squared | 0.031 | 0.034 | 0.023 | 0.026 |
| Control Variables | No | Yes | No | Yes |
| Time Trend | YES | YES | YES | YES |
| Province FE | YES | YES | YES | YES |
| Time FE | YES | YES | YES | YES |

Standard errors in parentheses, \*\*\* p<0.01, \*\* p<0.05, \* p<0.1

### 5.3 Different Effects in Different Regions

In order to further explore the regional heterogeneity of effect of the measures, we divide the sample into three groups: high, medium, and low according to the epidemic control capacity of the city (GDP per capita) and difficulty of epidemic control (population size). The sub-sample results are reported in Table 8. Panel A presents the results of subsample divided according to per capita GDP, and panel B presents the results subsample divided according to population size. We find that social distancing measures are effective in cities with high or low GDP; However, traffic control measures are more effective in regions with high GDP, the coefficient significance decreases successively in regions with medium and low GDP. The effect of traffic control measures is not ideal in regions with lower GDP. According to the section 2, we know that although traffic control measures effectively restrict the flow of population, it also makes it difficult to deliver supplies equipment and humanitarian aid to the affected areas, which controlling the spread of the epidemic while also increasing the difficulty of responding to the outbreak, so for cities of which control ability is originally lower (low GDP per capita), the practice of traffic control can’t obtain a good effect.

**Table 8.** Different Effects in Different Regions

| Panel A by GDP per capita |  |  |  |  |  |  |
| --- | --- | --- | --- | --- | --- | --- |
|  | (1) | (2) | (3) | (4) | (5) | (6) |
|  | Pergdp_H | Pergdp_M | Pergdp_L | Pergdp_H | Pergdp_M | Pergdp_L |
| Traffic | -0.0477*** | -0.0171** | -0.0127* |  |  |  |
| Control | (0.0165) | (0.0071) | (0.0072) |  |  |  |
| Social |  |  |  | -0.0456*** | -0.0287*** | -0.0315*** |
| Distancing |  |  |  | (0.0107) | (0.0045) | (0.0042) |
| Constant | -0.2360 | -0.7319** | 0.0179 | -0.3191 | -0.8175*** | -0.0133 |
|  | (0.6580) | (0.2944) | (0.2748) | (0.6571) | (0.2935) | (0.2731) |
| Observations | 4,275 | 4,140 | 4,140 | 4,275 | 4,140 | 4,140 |
| R-squared | 0.041 | 0.052 | 0.028 | 0.043 | 0.060 | 0.041 |
| Control | Yes | Yes | Yes | Yes | Yes | Yes |
| Variables |  |  |  |  |  |  |
| Time Trend | YES | YES | YES | YES | YES | YES |
| Province FE | YES | YES | YES | YES | YES | YES |
| Time FE | YES | YES | YES | YES | YES | YES |
| Panel B by population size |  |  |  |  |  |  |
|  | (1) | (2) | (3) | (4) | (5) | (6) |
|  | pop_H | pop_M | pop_L | pop_H | pop_M | pop_L |
| Traffic | -0.0389*** | -0.0247** | -0.0119 |  |  |  |
| Control | (0.0130) | (0.0125) | (0.0080) |  |  |  |
| Social |  |  |  | -0.0432*** | -0.0101 | 0.0067 |
| Distancing |  |  |  | (0.0074) | (0.0071) | (0.0049) |
| Constant | -0.4687** | 0.0500 | 0.2685 | -0.4903** | 0.0037 | 0.1843 |
|  | (0.2205) | (0.3085) | (0.3248) | (0.2203) | (0.3126) | (0.3248) |
| Observations | 7,510 | 2,835 | 2,210 | 7,510 | 2,835 | 2,210 |
| R-squared | 0.055 | 0.062 | 0.048 | 0.057 | 0.062 | 0.045 |
| Control | Yes | Yes | Yes | Yes | Yes | Yes |
| Variables |  |  |  |  |  |  |
| Time Trend | YES | YES | YES | YES | YES | YES |
| Province FE | YES | YES | YES | YES | YES | YES |
| Time FE | YES | YES | YES | YES | YES | YES |

Panel B presents the results of subsample divided according to population size. It can be seen that traffic control measures play a role in cities with a high population size and middle population sizes, while social distancing is only effective in cities with a high population size, while it is ineffective in cities with smaller population size. Measures, whether traffic control or social distancing, are aiming to reduce physical contact between people, and in cities with smaller populations, the contact there is already less than in high-density cities, so the effects of those measures fail in low-population cities. In short, the effects of the two measures are not the same in different regions, which depending on conditions of the city, and the implementation of prevention and control measures will incur huge social and economic costs, especially in the outbreak of COVID-19, in order to control the spread of the disease, many cities across the country have implemented strict prevention and control measures such as “lockdown” the city. While controlling the epidemic, they have also increased social and economic burdens. Therefore, when formulating an epidemic prevention and control policy, full consideration should be given to the local situation, implementing policies according to the city and achieving precise epidemic prevention.

### 5.4 Information Dissemination, Protection Awareness and Public Confidence

The effective control of the epidemic depends not only on the selection of prevention and control measures of the authority but also on the performance of the public. Therefore, in this section, we further use Baidu Search Index to explore the influence of the public’s access to information, protection awareness, and public confidence on the implementation effect of traffic control and social distancing measures. Fast and effective information dissemination can greatly reduce the information asymmetry of the public and to raise public awareness of the epidemic. We use the Baidu search index of “COVID-19” to characterize the degree of information spread among the public. A higher search index means that the public has learned about the novel coronavirus earlier and paid more attention to it; the public’s protection awareness and positive attitude will allow them to better carry out the measures, which can absolutely improve the effectiveness of policy implementation. Here, we use the Baidu search index of “the correct way of wearing a mask” to characterize protection awareness of the public. A higher search index indicates that people have a better sense of protection; we use Baidu search index of “Zhong Nanshan” to characterize people’s confidence about fighting the epidemic A high search index indicates that the public has more confidence to fight the epidemic, and it is even more convinced that the epidemic problem will be overcome.

We divide these three indexes into high, medium, and low, respectively, generate dummy variables, and construct the interaction terms between dummy variables and the measures. The results are shown in Table 9. Column (1) in Panel A constructs the interaction between traffic control and high information (H_information_H) and medium information (M_information_M), and column (2) constructs the interaction between social distancing and the two items; similarly, panel B presents the results of protection awareness, and panel C presents the results of public confidence. It can be seen that except for column (2) in Panel C, whether it is traffic control or social distancing measures, the coefficient of the interaction with high information dissemination (information_H), high protection awareness (awareness_H), and high public confidence (emotion_H) is significantly negative, that is, traffic control and social distancing measures have played a better role in cities with better information dissemination, high protection of the public,and strong confidence in epidemic prevention. It emphasizes the importance of paying attention to the public. While adopting traffic control and social distancing measures, the government should also increase the transparency of information disclosure, improve protection awareness of the public, and properly channel the public sentiment, strengthening public confidence.

**Table 9.**
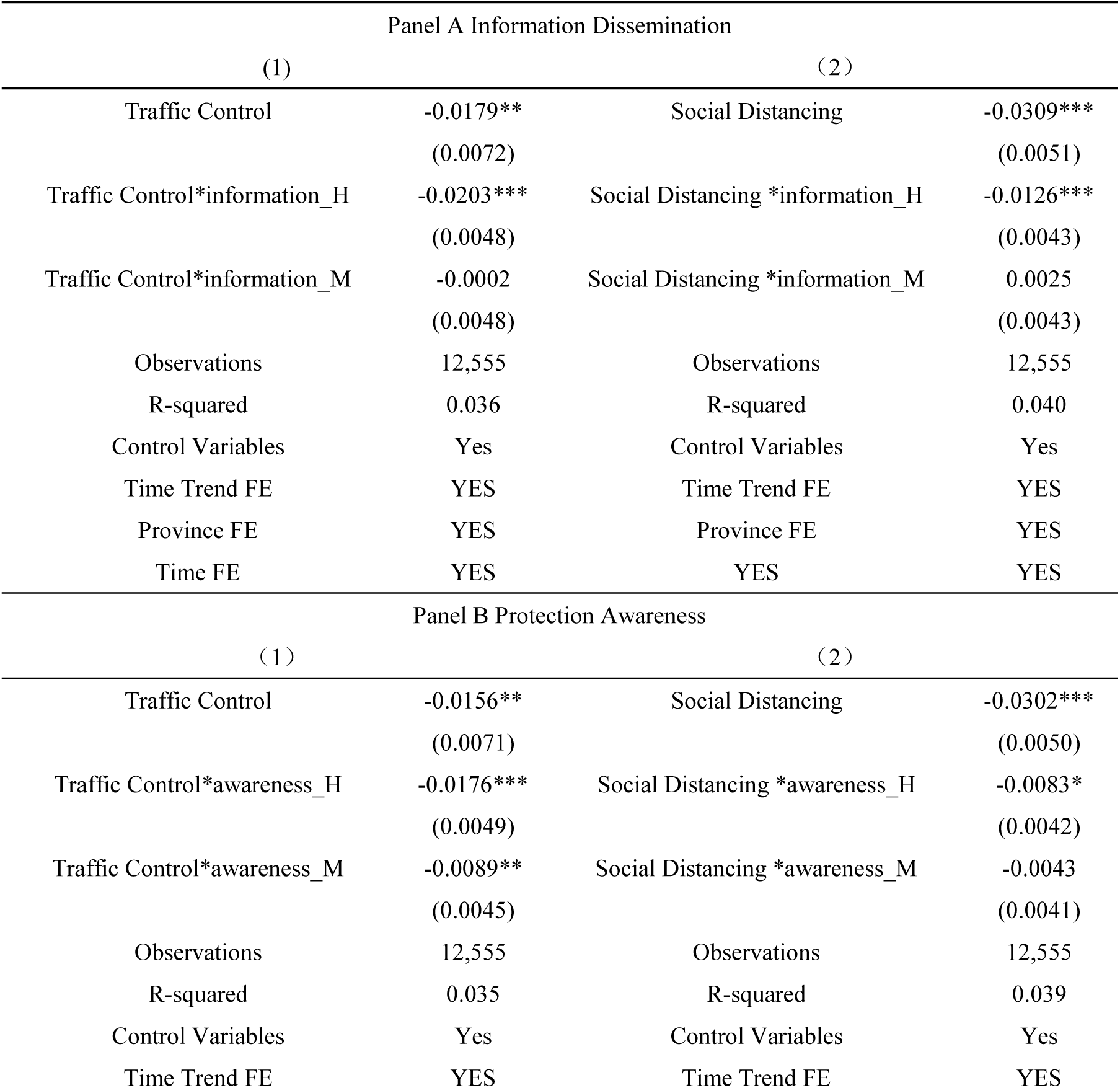
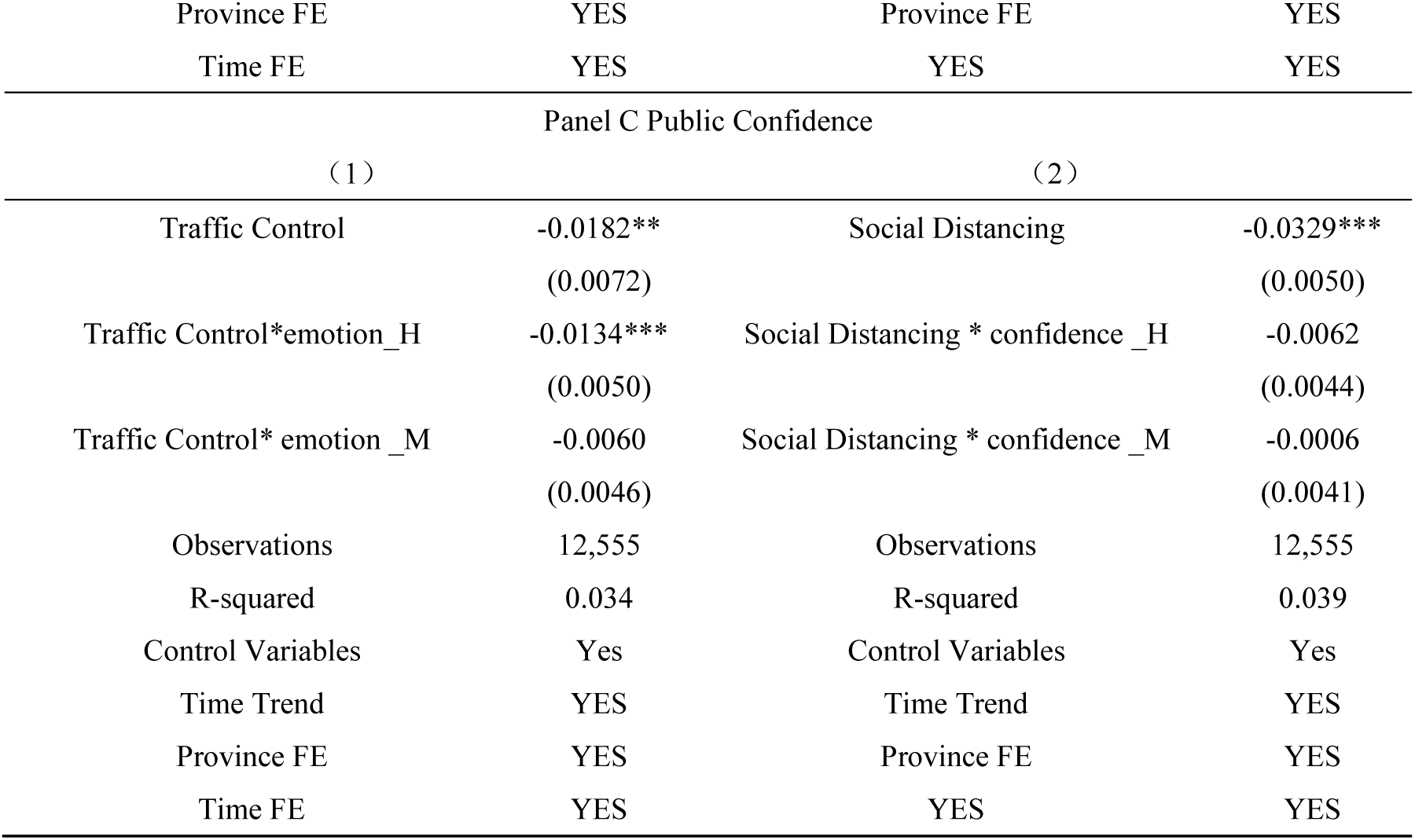
Information Dissemination, Protection Awareness and Public Confidence

## Conclusion and Discussion

The conclusions of this article are: ① In the prevention and control of the COVID-19 epidemic, traffic control and social distancing measures have played a very good role in controlling the spread of the epidemic. Nationwide, the effect of social distancing is better than traffic control. ② The two measures are complementary, and their combined effect will play a better role in epidemic prevention; ③ Traffic control and social distancing do not work everywhere. Traffic control only paly a role in cities with large GDP and population size, but not in cities with low GDP population size; In cities with lower population size, social distancing becomes inoperative. ④ The rapid and accurate transmission of information, a higher protection awareness of the public, and a stronger confidence of residents in epidemic prevention can promote the realization of the measure effects.

The conclusions above have important implications for public health policy. Considering the complementary relationship between the two measures, they should be coordinated with each other when making policies. However, considering the regional heterogeneity of their effect, it is necessary to formulate measures according to the city’s own situation, achieving precise epidemic prevention, and the need for such a strict measure as “lockdown” the city should be carefully weighed; considering the promotion of better information dissemination, high protection awareness and strong public confidence on the effect of the measures, we should increase the transparency of information, improve protection awareness of the public, guide emotions of the public in a proper way, enhancing public confidence.

Excessive prevention and control will inevitably bring negative effects. Severe traffic control completely blocked the flow of people and materials, which to some extent made it difficult for the timely replenishment of basic living needs. Social distancing measures such as closing communities and public places inevitably exacerbated the panic among the people and adversely affected the development of their mental health. Therefore, when actively taking prevention and control measures, we must also fully consider the possible negative impacts, reasonably grasp the implementation of the measures, and take supporting remedial measures to minimize the side effects of epidemic prevention and control.

## Data Availability

Some data, models, or code generated or used during the study are available from the corresponding author by request.

